# Early syphilis in Guangzhou, China: presentation, molecular detection of *Treponema pallidum*, and genomic sequences in clinical specimens and isolates obtained by rabbit infectivity testing

**DOI:** 10.1101/2023.10.17.23297169

**Authors:** Ligang Yang, Xiaohui Zhang, Wentao Chen, Arlene C. Seña, Heping Zheng, Yinbo Jiang, Peizhen Zhao, Rongyi Chen, Liuyuan Wang, Wujian ke, Juan C. Salazar, Jonathan B. Parr, Joseph D. Tucker, Kelly L. Hawley, Melissa J. Caimano, Christopher M. Hennelly, Farhang Aghakanian, Feifei Zhang, Jane S. Chen, M. Anthony Moody, Justin D. Radolf, Bin Yang

## Abstract

**Background:** The global resurgence of syphilis requires novel prevention strategies. Whole genome sequencing (WGS) of *Treponema pallidum* (*TPA*) using different specimen types is essential for vaccine development.

**Methods:** Patients with primary (PS) and secondary (SS) syphilis were recruited in Guangzhou, China. We collected ulcer exudates and blood from PS participants, and skin biopsies and blood from SS participants for *TPA polA* polymerase chain reaction (PCR); ulcer exudates and blood were also used to isolate *TPA* strains by rabbit infectivity testing (RIT). *TPA* WGS was performed on 52 ulcer exudates and biopsy specimens and 25 matched rabbit isolates.

**Results:** We enrolled 18 PS and 51 SS participants from December 2019 to March 2022. Among PS participants, *TPA* DNA was detected in 16 (89%) ulcer exudates and three (17%) blood specimens. Among SS participants, *TPA* DNA was detected in 50 (98%) skin biopsies and 27 (53%) blood specimens. *TP*A was isolated from 48 rabbits, with a 71% (12/17) success rate from ulcer exudates and 69% (36/52) from SS bloods. Twenty-three matched SS14 clade genomes were virtually identical, while two Nichols clade pairs had discordant *tprK* sequences. Forty-two of 52 unique *TPA* genomes clustered in an SS14 East Asia subgroup, while ten fell into two East Asian Nichols subgroups.

**Conclusions:** Our *TPA* detection rate was high from PS ulcer exudates and SS skin biopsies and over 50% from SS whole blood, with RIT isolation in over two-thirds of samples. Our results support the use of WGS from rabbit isolates to inform vaccine development.

**Summary:** We performed *Treponema pallidum* molecular detection and genome sequencing from multiple specimens collected from early syphilis patients and isolates obtained by rabbit inoculation. Our results support the use of whole genome sequencing from rabbit isolates to inform syphilis vaccine development.

## Introduction

During the past 20 years, syphilis, caused by the highly invasive and immunoevasive spirochete, *Treponema pallidum subspecies pallidum* (*TPA*), has resurged as a global public health problem [1–3]. In China, 57,196 new syphilis cases (12.7% of all cases in the country) were reported in Guangdong Province in 2021 [4]. Although the province has implemented syphilis intervention programs since 2000, the epidemic calls for novel prevention and control strategies, including vaccine development.

Darkfield microscopy (DFM) has been used traditionally for *TPA* direct detection from primary syphilis (PS) lesions, while detection of treponemal and non-treponemal antibodies in serum is the mainstay for syphilis diagnosis [5, 6]. Several studies have demonstrated the sensitivity of real-time *TPA* PCR on ulcer swabs and whole blood for diagnosis of PS and secondary syphilis (SS) [7–10]. The inability to cultivate *TPA* from clinical samples continues to hinder microbiologic diagnosis. Despite recent progress with *in vitro* cultivation [11–13], rabbit infectivity testing (RIT) by intratesticular inoculation of rabbits with clinical specimens remains the only means of recovering *TPA* strains [14, 15].

Whole-genome sequencing (WGS) of *TPA* genomes from geographically diverse regions is essential for development of a syphilis vaccine with global efficacy. At present, most *TPA* sequences have originated from Europe, North America and Australia; fewer strains have been identified from Asia [16–19]. The majority of clinical specimens used for WGS have been PS ulcer swabs with high *TPA* burdens [19–22]. Skin biopsies and whole blood from SS patients are other potential specimens but have considerably lower *TPA* burdens than ulcer exudates [5, 10, 23–25]. RIT allows strain amplification from clinical specimens with low *TPA* burdens, but it is unclear whether the genomes of treponemes obtained by rabbit infectivity testing accurately reflect those in the corresponding clinical specimens.

In this study, we recruited patients with PS and SS from Guangzhou, China as part of a parent study for syphilis vaccine development [26]. We collected ulcer exudates and whole blood specimens from PS patients, and skin biopsy and whole blood specimens from SS patients for quantitative PCR (qPCR) and RIT. WGS was performed on clinical specimens with sufficient *TPA* DNA copy numbers and on corresponding rabbit isolated strains. In addition to characterizing the clinical presentation of PS and SS in our patient population, we aimed to (i) compare *TPA* qPCR results and RIT between specimen types, (ii) assess the effect of rabbit passage on the genomic sequences of *TPA* strains isolated from patients, and (iii) phylogenetically analyze *TPA* strains by WGS as a prelude for syphilis vaccine development.

## Methods

### Ethics statements

This research was approved by the Medical Ethics Committee Dermatology Hospital of Southern Medical University (GDDHLS-20181202). Rabbit experimentation was approved by the Ethics Committee of Dermatology Hospital of Southern Medical University (approval no. GDDHLS-20181202,12/12/2018) and South China Agricultural University Experimental Animal Ethics Committee (approval no. 2020C004, 07/05/2020). All rabbits were housed under approved biosafety conditions at South China Agricultural University. Their diet, care, and maintenance conformed to institutional regulations.

### Patient recruitment

A convenience sample of patients visiting the STI department of the Dermatology Hospital of Southern Medical University (DHSMU) from December 2019 to March 2022 were recruited for the study. Individuals aged ≥ 18 years and diagnosed with PS or SS were eligible for enrollment [26]. Diagnosis of PS was based on the presence of one or more typical anogenital ulcers with a positive DFM and/or reactive nontreponemal (Toluidine red unheated serum test, TRUST) and treponemal (*Treponema pallidum* particle agglutination, TPPA) tests [5]. Diagnosis of SS was based on characteristic skin or mucosal lesions plus reactive TRUST and TPPA tests [5, 27].

All study participants received standard care including physical examination and treatment of STIs as indicated. For treatment of PS and SS, we provided 2.4 million units (MU) of intramuscular benzathine penicillin G (BPG) weekly for 2 consecutive weeks (total of 4.8 MU) in accordance with Chinese Center for Disease Control and Prevention treatment guidelines [27]. Patients were asked to return within 90 days for a follow-up examination and repeat TRUST titers; TPPA testing was repeated if the initial test was negative.

### Data and specimen collection

A case report form (CRF) was used to collect demographic and clinical data, including sexual orientation, sexual and medical histories [26]. Examination and laboratory findings were also recorded in the CRF. For PS patients, we recorded the number and location of ulcers. For SS patients, we recorded the type and location of rash, as well as other mucocutaneous manifestations. For rashes, we estimated Body Surface Area of Involvement (BSA) scores, a disease outcome predictor used in other skin diseases, such as psoriasis [28].

From PS participants, we collected ulcer exudates for DFM and research testing. The first swab was used for DFM, and the residual of the first swab plus a second swab were placed into 1 mL of TpCM-2 medium [13] for RIT [15]. The third and fourth swabs were placed in a 1.5 mL tube containing DNA/RNA shield (Zymo Research, Irvine, CA, USA; R1100–250) for subsequent DNA extraction. Whole blood specimens were also collected from PS participants for DNA extraction. From SS participants, two 4 mm punch skin biopsies were obtained from rashes and stored in tubes containing DNA/RNA shield. Whole blood specimens were also collected from SS participants for RIT and DNA extraction.

### Assessment of T. pallidum burdens by TaqMan qPCR

DNA was extracted from participant samples using the DNeasy Blood & Tissue Kit (QIAGEN, Hilden, Germany) according to manufacturer’s instructions; 100 μL of nuclease-free water was used to elute the DNA, followed by −80 °C storage. TaqMan PCR for *TPA polA* was performed as previously described by Chen *et. al.* [29] using 1.25 μL of 10 μM primers, and measured on Real-Time PCR Instruments (BioRad, Hercules, CA, USA)

### Rabbit-infectivity testing

RIT was conducted as previously described [15]. In short, adult New Zealand White rabbits (males, 3 months of age, 2.5-3 kg) were pre-screened to confirm non-reactive syphilis serologies at baseline. Genital ulcer exudates from participants diluted with 1 mL of TpCM-2 medium or 1ml of freshly collected whole blood was injected into each testis. The serologic status of the rabbits was monitored weekly by *TPA* particle agglutination (TPPA) beginning with the second month after inoculation. Seroconverted rabbits were sacrificed, and their testes were aseptically removed for *TPA* isolation. Seronegative rabbits were euthanized after 3 months followed by passage of testicular extract to a second rabbit.

### Whole genome sequencing and bioinformatic analysis

WGS was carried out on ulcer exudates from individuals with PS, skin biopsy specimens from SS patients, and rabbit-passaged isolates using custom 120-nucleotide RNA oligonucleotide baits obtained from Agilent Technologies (Santa Clara, CA, USA). *TPA* enrichment was performed using the Sure Select XT Low Input kit at the University of North Carolina at Chapel Hill (UNC) and the Sure Select XTHS2 kit at SMU, following the previously established protocol [16]. The resulting *TPA*-enriched libraries were sequenced using the Illumina MiSeq platform at UNC or the NovaSeq platform at SMU, generating paired-end, 150 bp reads. The raw sequencing data, with residual human reads removed, has been deposited at the Sequence Read Archive database (PRJNA815321). Fifty-two *TPA* genomes from China included in this manuscript are also reported in separate study by Seña *et al*. [26].

The sequencing reads and publicly available data were processed using a conservative bioinformatic pipeline with minor modifications [16], which is available at https://github.com/IDEELResearch/TPAllidum_genomics. Only sequences with at least 80% of their genome covered 3x were retained for variant calling. Variant calling was performed with GATK HaplotypeCaller [30], using joint genotyping using GenotypeVCFs module and hard filtering by VariantFiltration.

Comparison of paired *TPA* genomes (rabbit-passaged versus direct sequencing from clinical samples) was conducted manually using a VCF file obtained from joint genotyping, applying strict filtering parameters (depth ≥ 8x, genotype quality ≥ 20, and variant allele frequency (VAF) ≥ 0.85). Variants genes (*e.g., tprK*) were considered to be heterogeneous if portion of reads supporting less frequent alleles were greater than 15% (*i.e.,* VAF between 0.15 and 0.85). To generate a multiple alignment using the MAFFT (v7.520) [31], we combined 52 Nichols- and SS14-clade consensus sequences generated in this study, 109 published *TPA* genomes and 2 outgroup genomes (*Treponema subsp. endemicum* Bosnia A and *subsp. pertenue* Samoa D). The resulting alignment was then inputted into Gubbins (v3.2.1),[32] which processed the alignment with default parameters for predicting and masking homologous recombination regions. Phylogenetic tree was constructed under the GTRGAMMA model with 1000 bootstraps replicates by Gubbins (v3.2.1), which then was visualized using ggtree [33]. The genotypic resistance of macrolide was analyzed by competitive mapping as previously described by Beale *et al.* [18, 34].

### Statistical analysis

Descriptive analyses were conducted on participants’ demographics and clinical characteristics, *TPA* PCR, and RIT results. All data were analyzed using SPSS 22.0 (IBM® SPSS® Statistics).

## Results

### Sociodemographic data, clinical characteristics, and laboratory findings

From December 2019 to March 2022, 154 participants met the screening criteria (Figure 1). Of the 115 eligible persons, 69 individuals (18 PS and 51 SS) consented for enrollment. Among enrolled participants, the median age was 27 (IQR: 22-32); 84% were male, and 33% were men who have sex with men (MSM) or men who have sex with both men and women (MSMW) (Table 1). All participants but one were HIV-negative, 9% had a history of prior syphilis, and 22% had a history of other STIs.

**Figure 1.**
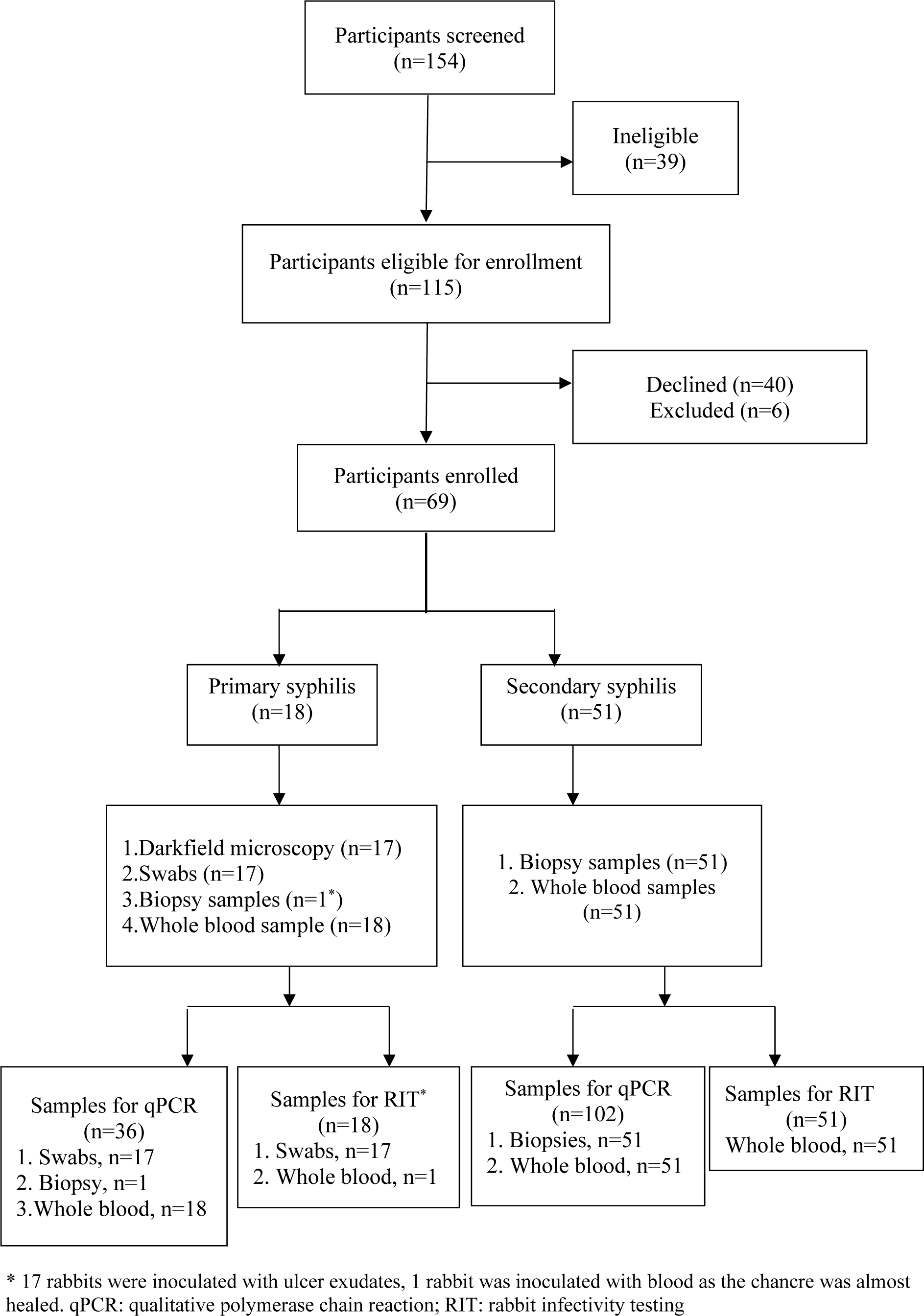
Screening and enrollment algorithm of participants with primary and secondary syphilis.

**Table 1.** Sociodemographic Characteristics of 69 Patients Enrolled in Guangzhou, China.

| Characteristics | Primary syphilis<br>n=18 (%) | Secondary<br>syphilis n=51<br>(%) | Total<br>n=69 (%) |
| --- | --- | --- | --- |
| Age (median/inter-quartile range) | 27 (IQR 23-31) | 27 (IQR 22-33) | 27 (IQR 22-32) |
| Gender |  |  |  |
| Women | 1 (6) | 10 (20) | 11 (16) |
| Men | 17 (94) | 41 (80) | 58 (84) |
| Sexual orientation |  |  |  |
| Heterosexual | 12 (67) | 33 (65) | 45 (65) |
| Men who have sex with men | 4 (22) | 9 (18) | 13 (19) |
| Men who have sex with men and women | 2 (11) | 8 (16) | 10 (14) |
| Declined to answer | 0 (0) | 1 (2) | 1 (1) |
| Marital status |  |  |  |
| Single | 12 (67) | 34 (67) | 46 (67) |
| Married | 6 (33) | 12 (24) | 18 (26) |
| Divorced/separated/widowed | 0 (0) | 5 (10) | 5 (7) |
| Educational status |  |  |  |
| Middle school or less | 1 (6) | 6 (12) | 7 (10) |
| High school | 2 (11) | 15 (29) | 17 (25) |
| Trade school | 6 (33) | 12 (24) | 18 (26) |
| Undergraduate or Graduate | 9 (50) | 18 (35) | 27 (39) |
| Age of first sexual encounter |  |  |  |
| 15~17 years | 2 (11) | 13 (25) | 15 (22) |
| ≥18 years | 16 (89) | 38 (75) | 54 (78) |
| Reported sexual partners in past 3 months |  |  |  |
| None | 0 (0) | 5 (10) | 5 (7) |
| One | 9 (50) | 25 (49) | 34 (49) |
| Two | 3 (17) | 9 (18) | 12 (17) |
| Three or more | 6 (33) | 12 (24) | 18 (26) |
| Reported sexual partners in past 12 months |  |  |  |
| None | 0 (0) | 1 (2) | 1 (1) |
| One | 4 (22) | 8 (16) | 12 (17) |
| Two | 3 (17) | 14 (27) | 17 (25) |
| Three or more | 11 (61) | 28 (55) | 39 (57) |
| Commercial sex engagement | 2 (11) | 11(22) | 13 (19) |
| HIV status at enrollment |  |  |  |
| Positive | 0 (0) | 1 (2) | 1 (1) |
| Negative | 18 (100) | 50 (98) | 68(99) |
| History of syphilis | 1 (6) | 5 (10) | 6 (9) |
| History of other sexually transmitted infections | 4 (22) | 11 (22) | 15 (22) |

Seventeen of the 18 participants with PS were male, with the majority (61%) presenting with solitary ulcers (Table 2, Figure 2A and 2B); one male participant presented with a nearly healed chancre. Spirochetes were visualized by DFM in 12 of the 17 (71%) patients with exudative lesions. The TRUST and TPPA tests were positive in 8 (45%) and 16 (89%) PS patients, respectively. *TPA* DNA was detected in 16 of 18 (89%) specimens collected from chancres and 3 of 18 (17%) PS blood specimens. *TPA polA* copy numbers ranged from 1.43 – 2250 copies/μl in swabs and 0.288- 37.99 copies/μl in whole blood (Supplemental Table 1). All 12 DFM positive specimens were positive by *TPA* PCR (Supplemental Table 1). *TPA* DNA was detected in ulcer exudates and whole blood in two PS cases with negative TRUST and TPPA results (Supplemental Table 1).

**Figure 2.**
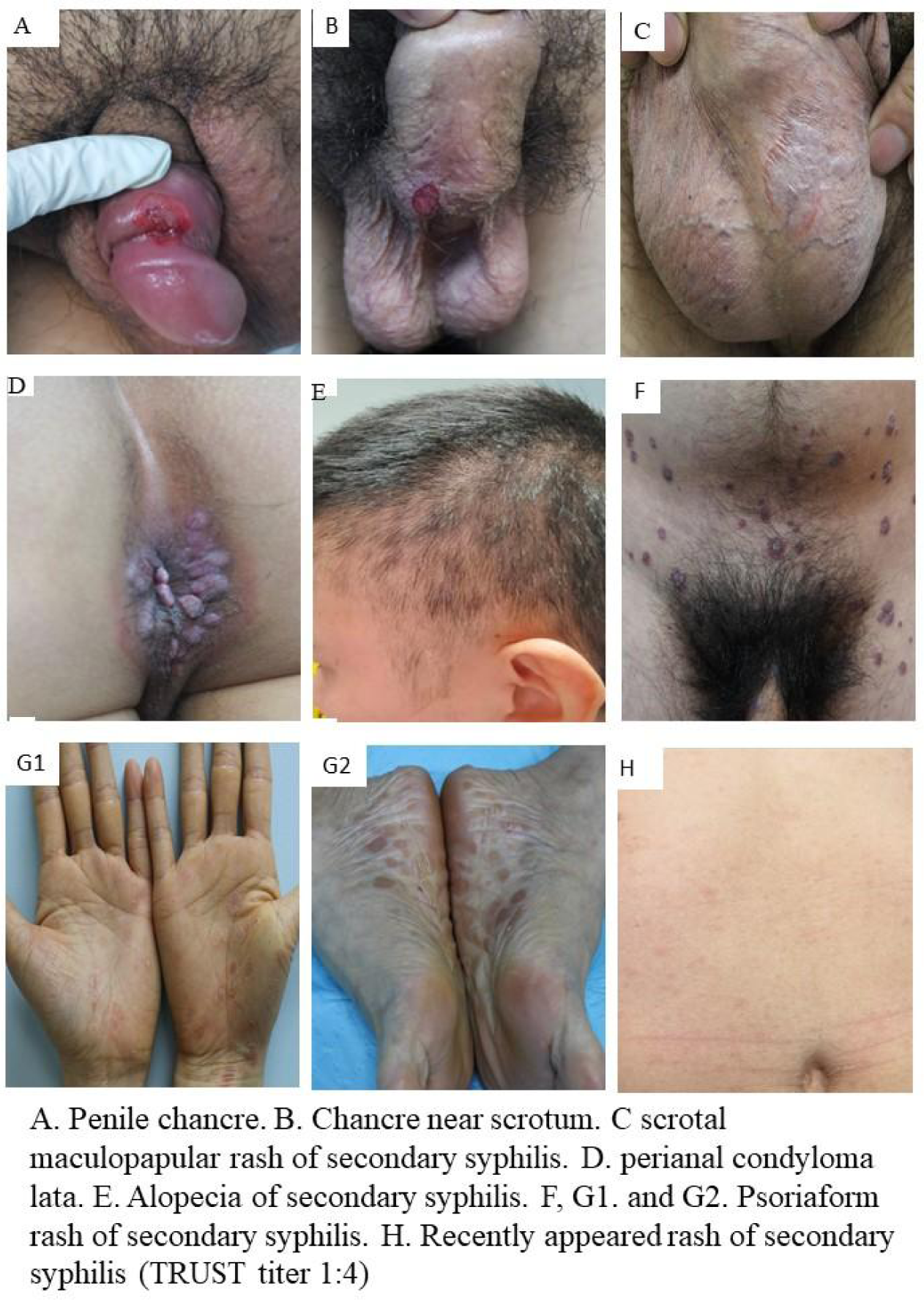
Representative clinical manifestations in study participants

**Table 2.** Clinical characteristics and laboratory findings for patients with primary syphilis.

| **Characteristic** | **Number**<br>**n=18 (%)** |
| --- | --- |
| Location of genital ulcers |  |
| Vulva | 1 (6) |
| Penis | 17 (94) |
| Number of genital ulcers |  |
| 1 ulcer | 11 (61) |
| 2 ulcers | 2 (11) |
| 3 or more ulcers | 5 (28) |
| Darkfield microscopy<sup>a</sup> |  |
| Positive | 12 (71) |
| Negative | 5 (29) |
| TPPA |  |
| Positive | 16 (89) |
| Negative | 2 (11) |
| TRUST titer |  |
| ≥1:64 | 1 (6) |
| 1:16 - 1:32 | 2 (11) |
| 1:1 - 1:8 | 5 (28) |
| Negative | 9 (50) |
| Unknown | 1 (6) |
| TPPA+TRUST |  |
| Both positive | 9 (50) |
| Both negative | 2 (11) |
| qPCR chancres<sup>b</sup> |  |
| Positive | 16 (89) |
| Negative | 2 (11) |
| qPCR blood |  |
| Positive | 3 (17) |
| Negative | 15 (83) |

<sup>a</sup> n=17; one patient did not undergo dark field microscopy because the ulcer was nearly healed.
<sup>b</sup>qPCR was performed on 17 swabs and one biopsy.

Forty two of the 51 (82%) SS participants presented with a rash only; the other nine presented with other manifestations, such as condyloma lata and alopecia (Figure 2 and Table 3). BSA scores were <3% in 15 participants (30%), 3-10% in 10 (19%), and >10% in26 (51%). All SS participants had reactive TRUST and TPPA tests. *TPA* DNA was detected in 50 of 51 (98%) skin biopsies and 27 of 51 (53%) whole blood specimens. *TPA polA* copy numbers ranged from 0.610 - 10,200 copies/μl in skin biopsies and 0.110 - 3,850 copies/μl in blood (Supplemental Table 2).

**Table 3.** Clinical characteristics and laboratory findings among patients with secondary syphilis.

| **Characteristic** | **Number**<br>**n= 51 (%)** |
| --- | --- |
| Manifestations |  |
| Rash only | 42 (82) |
| Rash and condyloma lata | 1 (2) |
| Rash and mucous patches | 1 (2) |
| Rash and alopecia | 6 (12) |
| Condyloma lata only | 1 (2) |
| Location of rash |  |
| Trunk | 39 (76) |
| Extremities | 33 (65) |
| Palms and soles | 23 (45) |
| Ano-genital area | 15 (29) |
| Face | 2 (4) |
| BSA score for rash |  |
| <3%, mild | 15 (30) |
| 3%~10%, moderate | 10 (19) |
| >10%, severe | 26 (51) |
| Initial TRUST titer |  |
| ≥ 1:64 | 21 (41) |
| 1:16 - 1:32 | 28 (55) |
| 1:4 - 1:8 | 2 (4) |
| <1:4 | 0 |
| *TPA* PCR of skin biopsy |  |
| Positive | 50 (98) |
| Negative | 1 (2) |
| *TPA* PCR of blood |  |
| Positive | 27 (53) |
| Negative | 24 (47) |

BSA: Body Surface Area.

### Rabbit infectivity testing

We inoculated New Zealand white rabbits with 17 ulcer exudates and 52 whole blood specimens (one from the PS patient with a nearly healed chancre and 51 from SS patients; Table 4). Fifty-one rabbits (74%) developed reactive TPPA tests, which included 14/17 (82%) rabbits inoculated with ulcer exudates and 37/52 (71%) inoculated with blood. The median time to TPPA positivity following inoculation was 42 days for ulcer exudates versus 50 days for whole blood. *TPA* strains were isolated from 48 of the 51 rabbits that TPPA seroconverted – 12/17 (71%) from ulcer exudates and 36/52 (69%) from blood (Table 4). One of two rabbits inoculated with PCR-negative exudates and 16 of 25 rabbits inoculated with PCR-negative blood samples also yielded isolates (Table 4). Conversely, RIT was negative in two of 15 PCR-positive ulcer exudates and seven of 27 PCR-positive blood specimens. *TPA polA* copy numbers in the two PCR-positive RIT-negative ulcer exudates were comparable to those in PCR-positive RIT-positive samples (Suppl. Table 1). In contrast, *polA* copy numbers were at the low end in five of the seven PCR-positive RIT-negative blood samples (Suppl. Table 2).

**Table 4.** Comparison of *TPA* qPCR and rabbit infectivity testing (RIT) from 69 participants.

|  | **RIT (-)**<br>**(%)** | **TPPA**<br>**seroconverted (%)** | ***TPA* isolated**<br>**(%)** |
| --- | --- | --- | --- |
| **Ulcer specimens**<br>(n=17) |  |  |  |
| qPCR+, n=15 | 2 /15 (13) | 13/15 (87) | 11/15 (73) |
| qPCR-, n=2 | 1/2 (50) | 1/2 (50) | 1 /2(50) |
| Total | 3/17 (18) | 14/17 (82) | 12/17 (71) |
| **Blood specimens<sup>#</sup>**<br>(n=52) |  |  |  |
| qPCR+, n=27 | 7/27 (26) | 20/27 (74) | 20/27 (74) |
| qPCR-, n=25 | 8/25 (32) | 17/25 (68) | 16/25 (64) |
| Total | 15/52 (29) | 37/52 (71) | 36/52 (69) |

<sup>#</sup>51 from SS patients and 1 from a patient with a nearly healed chancre.

*Comparison of TPA genomic sequences from paired clinical samples and rabbit isolates* We successfully generated a comprehensive set of 77 *TPA* genomes from 69 participants. This dataset included 25 paired genomes, comprising clinical samples and their corresponding rabbit-passaged isolates, as well as 27 unmatched genomes consisting of 23 rabbit-passaged isolates and 4 clinical samples. Supplementary Table 3 presents detailed information regarding the samples subjected to WGS.

We compared *TPA* genomes from 25 clinical specimens and corresponding rabbit isolated strains to ascertain whether WGS from rabbit-passaged isolates accurately reflect those in the source clinical specimens. To ensure the reliability and rigor of the results, we only included variants in the analysis that passed conservative filtering criteria.

Twenty three of the 25 genomes were identified as SS14-lineage, while the remaining two were Nichols-like. Within the SS14-like group, we observed 6 discordant variants at 3 specific genomic sites when we applied the rigorous variant filtering criteria (Supplementary table 4). However, with manual inspection, we found three additional discrepant heterogenous variants (including 1 SNP and 2 indels) in *tprK* (Supplementary table 5). Within the two paired Nichols-lineage genomes, we identified a total of 28 discrepant variations. Two of these discordant variants were found in one of the paired genomes, while the remaining 26 discordant variants were present in the other (Supplementary Table 6). All 28 discordant variants were found exclusively within the *tprK* gene (Supplemental Table 6). Due to the complexity of the *tprK* region and the limitations of short-read sequencing, we are unable to go deeper to potentially uncover additional *tprK* variants. Overall, the results clearly showed that rabbit passage has minimal effects on *TPA* genomic stability.

### Phylogenetic diversity of new TPA strains

We constructed a phylogenetic tree to assess the diversity of the newly sequenced *TPA* strains. For genomes from matched samples and rabbit isolates, we prioritized the use of clinical samples. When clinical samples were unavailable, we utilized genomes from rabbit-passaged isolates. The final analyzed dataset, therefore, consisted of 29 genomes directly sequenced from clinical samples and 23 genomes from rabbit-passaged strains (7 from ulcer exudates and 16 from blood samples). We also included 109 previously published *TPA* genomes, including 28 *TPA* strains from Japan and 22 others available from China up to the present date. Additionally, our analysis included outgroup genomes from *Treponema pallidum* subsp. *endemicum* Bosnia A and *Treponema pallidum* subsp. *pertenue* Samoa D (Figure 3). The 52 newly sequenced Chinese strains fell into three subgroups: 42 clustered in the SS14-Omega East Asia subgroup, eight were classified into the Nichols C subgroup, and two were assigned to the Nichols E subgroup.

**Figure 3.**
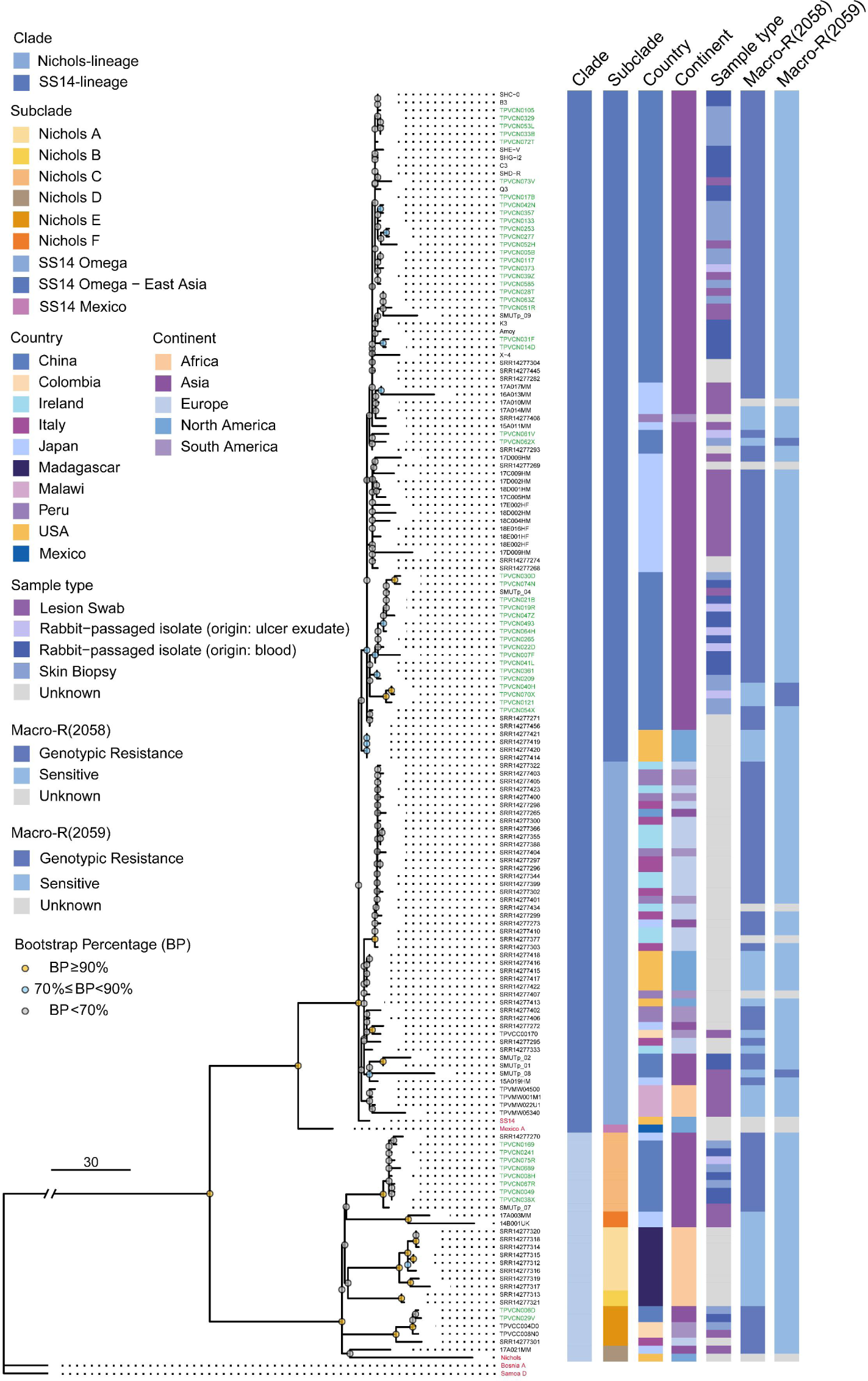
Phylogenetic diversity of new Chinese and published *Treponema pallidum* strains. The recombination-masked phylogeny tree was constructed using genetic sequences from 52 newly identified Chinese strains, 109 previously published strains [16, 17, 22, 52, 53, 55], and 2 outgroup strains (TPE and TEN) for comparison. The new Chinese *TPA* strains are highlighted in green on the tree, while five reference strains (SS14, Nichols, Mexico A, TPE, and TEN) are designated in red.

*TPA* strains harboring mutations in the 23S rRNA gene at positions A2058G or A2059G are associated with genotypic resistance to macrolides [34, 35]. Forty-eight of the newly sequenced Chinese *TPA* strains exhibited genotypic macrolide resistance, predominantly through the A2058G allele mutation, while four strains were harbored genotypic macrolide resistance with A2059 allele mutation (**Figure 3**).

## Discussion

We enrolled participants who presented with a wide range of PS and SS clinical manifestations, allowing for multiple specimen types for *TPA* molecular detection and genome sequencing. In addition, we were able to adapt a traditional, highly specialized syphilis research tool, RIT, for an entirely novel purpose of direct relevance to vaccine development – obtaining isolates for WGS. The *TPA* genomic sequences from our site with accompanying clinical data have provided new insights into the syphilis epidemic in Guangzhou, paving the way for molecular epidemiological studies that can synergize with future vaccine strategies.

We observed several interesting clinical findings in our study. Thirty-nine percent of our PS participants presented with multiple ulcers in the anogenital area although single ulcers are typical [36, 37]; over half of all PS participants had negative TRUST titers, underscoring the importance of direct detection for diagnosis. We found the sensitivity of DFM for PS in our study to be 71%, compared to the range of 70-100% in the literature [10, 38, 39]. Among our SS participants, we noted that over half of the SS participants had >10% BSA scores suggesting widespread dermal involvement from *TPA*. Rashes on the palms and soles are often indicative of a secondary syphilis diagnosis, reported in 40% to 80% of cases in the literature [40, 41]; however, less than half of our participants had these typical findings.

We found that the sensitivity of *TPA polA* PCR was 89% for chancres and 17% for whole blood from PS participants, compared to 98% for skin biopsies and 53% for blood from SS participants. Although our PCR sensitivity for whole blood was higher than in some reports [10, 42, 43], the overall low detection rates are likely explained by the low concentration of live *TPA* circulating in blood and the small volumes of specimens used for PCR testing. We used PCR to detect *TPA* in SS biopsy specimens and achieved an extraordinarily high positivity rate, supporting its use for diagnosis if it can be performed.

Our results support the importance of DFM and PCR to assist in the diagnosis and staging of syphilis [5, 10, 43].

RIT has long been regarded as the gold standard for detection of small numbers of live treponemes in clinical specimens [14, 15]. While a variety of specimens have been successfully inoculated into rabbits over the years [14, 15], studies of acquired syphilis in the antibiotic era have focused on cerebrospinal fluid to identify patients with asymptomatic neurosyphilis [44, 45]. Our study, therefore, is unique in its emphasis on RIT of ulcer exudates and whole blood. Interestingly, we isolated *TPA* in some rabbits inoculated with PCR-negative blood, emphasizing the extraordinary sensitivity of RIT. The differences in sensitivity between RIT and PCR may reflect the volume of whole blood used for RIT versus *TPA* qPCR. On the other hand, we also noted that specimens with detectable *TPA* DNA were not uniformly RIT-positive. Treponemal burdens in samples at or near the minimal infectious inoculum of individual *TPA* strains may partially explain these discordant results [46], while the presence of *TPA* DNA does not always reflect viable treponemes [47]. Moreover, it is not widely appreciated that clinical strains do not readily adapt to rabbits [48]. Indeed, as shown herein, amplification of the initial inoculum by serial rabbit passage is typically required. We also noted that organisms may not be recoverable from rabbits that clearly became infected based on conversion of their TPPA tests.

A critical question in the use of RIT for WGS is whether rabbit passage induces genomic alterations in the parental strain harbored by the inoculated clinical sample. We addressed this issue by performing the first large scale comparison of genomic sequences from matched clinical samples and rabbit isolates. The results clearly showed that rabbit passage has minimal effects on *TPA* genomic stability. The genomes of all 23 recovered SS14 clade strains were essentially identical to their clinical counterparts, while variation in the two Nichols clade isolates was confined to their *tprK* loci. Evidence from WGS for recombination in *TPA* strains [49, 50] implies that co-infection must occasionally occur, presumably in genital ulcers where high treponemal burdens would maximize the probability for genetic exchange. However, in all 25 matched cases, genomes in clinical samples and rabbit isolates clearly derived from the same *TPA* strain. We anticipate that future comparisons of this type will eventually identify co-infected individuals.

The newly sequenced *TPA* genomes were found to cluster within the SS14-Omega Asia, Nichols C, and E subgroups. Interestingly, although three previously reported Chinese SS14-clade *TPA* strains were found to cluster in the SS14-Omega subgroup [16], the vast majority of Chinese *TPA* strains published to date belonged to the SS14-Omega East Asian subgroup .[51–53]. Furthermore, the Chinese *TPA* strains showed a high degree of relatedness to publicly available Japanese *TPA* strains, both in the SS14-lineage and Nichols-lineage strains. Notably, genotypic resistance to macrolides was observed in all newly sequenced strains, indicating that azithromycin should not be considered as an alternative drug for treating syphilis patients in China.

There were several limitations in our study which included our small sample size enrolled from one site in Guangzhou. However, we encountered challenges with the COVID-19 pandemic in China, and our population is representative of patients presenting to a provincial hospital with a broad catchment area. We only inoculated one whole blood specimen from a PS participant who had a healed ulcer, therefore limiting our assessment of this sample type for RIT isolation. Based on the low *TPA* copy numbers in blood during PS, however, we would not expect similar RIT isolation rates to whole blood from SS participants. Our short-read sequencing and analysis approach prevents us from fully resolving challenging but important loci such as *tprK*. However, our conservative variant calling approach reduced ambiguity and allowed us to focus our analysis on high-confidence discordant loci.

Our study serves as a reminder of the remarkable diversity of PS and SS clinical manifestations, and the importance of sensitive detection methods. We showed that DFM, which is no longer performed in most STI clinics, remains a valuable tool for rapid diagnosis of exudative mucocutaneous lesions. We also confirmed previously reported high qPCR sensitivities for ulcer exudates and skin biopsies. Although exclusively a research tool, RIT provided a powerful means to obtain *TPA* strains for WGS from blood – a compartment critical to the systemic disease process but heretofore inaccessible to genomic investigation. The greater availability of WGS may open the door to genome-based studies of local sexual networks that can assist in the development of novel epidemiologic strategies for syphilis control in hyper-endemic areas. *TPA* outer membrane proteins are widely considered the prime candidates for syphilis vaccine design [54]. Identification of immunologically relevant outer membrane protein variants through WGS of *TPA* strains from clinical specimens and RIT passaged isolates will greatly facilitate development of a globally efficacious syphilis vaccine.

## Contributors

ACS, JCS, KH conceptualized the overall project and the study, and HZ, JBP led genomic sequencing efforts. All authors contributed to study design and implementation. LY and BY led the clinical sites in enrollment, participant data and specimen collection. Both contributed equally to this activity. XZ, RC, LW and WK participant in participant enrollment, data and specimen collection. XZ, PZ FZ and JSC conducted data curation and formal analysis of the clinical data. WC conducted data curation and formal analysis of the genomic data. ACS, LY, JSC, JDT, JCS developed the case report forms. FZ assisted with participant enrollment. WC, YJ, HZ conducted the rabbit-infectivity testing. WC anc CMH conducted the DNA extraction and *polA* PCR quantitation. FA, WC, HZ, CMH, MJC, JBP contributed to the genome sequencing and phylogenetic analyses. LY, XZ, WC, ACS, JDR, JSC prepared the tables and figures. XZ, PZ, FZ, MJC, ACS, JCS provided input into data analyses. JDT, HZ, BY, JCS, JDR, JBP provided administration, resources and supervision for the clinical or research laboratory sites. ACS, JDR, MAM contributed to the funding acquisition from the NIH and BMGF. ACS, JSC, FA, and JBP verified the underlying data. LY, XZ, WC prepared the manuscript, which was reviewed and edited by all authors. All authors had full access to all the data in the study and had final responsibility for the decision to submit for publication.

## Acknowledgments

This project was funded by the US National Institutes of Health National Institute for Allergy and Infectious Disease (NIAID) (U19AI144177 to JDR and MAM). This work also was supported, in part, by the Bill & Melinda Gates Foundation (INV-036560 to ACS), strategic research dollars from Connecticut Children’s, NIAID (T32 T32AI007151 to FFA), the National Nature Science Foundation of China (82220108006 to BY), the National Institute of Virology and Bacteriology (Programme EXCELES, ID Project No. LX22NPO5103, Funded by the European Union - Next Generation EU to DS). Under the grant conditions of the Foundation, a Creative Commons Attribution 4.0 Generic License has already been assigned to the Author Accepted Manuscript version that might arise from this submission. Fifty-two *TPA* genomes from China included in this manuscript are also reported in separate study by Seña et al. [26].

The authors would like to thank all of the study participants and research staff in Guangzhou, Cali, Lilongwe, and Chapel Hill. They would like to thank Myron Cohen for his support and guidance as a prior member of the Scientific Review Committee for the grant. They would also like to thank Andreea Waltmann and Fredrick Nindo for early contributions to genomic sequencing methods development and data analysis pipelines, respectively.

## Data availability

The deidentified participant REDCap database will be made publicly available at the timeof publication. Requests to share the informed consent forms and database can be made by emailing the corresponding authors at. Raw sequencing data from this study with residual human reads removed are available through the Sequence Read Archive (SRA, BioProject PRJNA815321). Data supporting the findings of this study are available within the manuscript and Supplementary material.

